# Influence of Built Environment on Bike Sharing Usage under COVID-19

**DOI:** 10.1101/2021.09.17.21263721

**Authors:** Hongtai Yang, Zishuo Guo, Jinghai Huo, Linchuan Yang

## Abstract

Bike sharing, as an important component of urban public transportation, has played a more important role during the COVID-19 pandemic because users could ride bikes in open space and avoid the risk of infection. Leveraging the trip data of the Divvy bike sharing system in Chicago, this study sets to explore the change of ridership that COVID-19 has brought and the built environment factors that influence the spatial variation of ridership under the pandemic. Results show that the ridership declines by xx% in total. To account for the spatially heterogeneous relationship between the built environment and the ridership, the geographically weighted regression (GWR) model and the semi-parametric GWR (S-GWR) model are constructed. By comparing the model results, we find that the S-GWR model outperforms the GWR and the multiple linear regression model. The results of the S-GWR model indicates that education employment density, distance to subway, COVID-19 cases and ridership before COVID-19 are global variables. The ridership between residential density, office employment density and the ridership vary across space. The results of this study could provide useful reference to transportation planners and bike sharing operators to determine the high bike sharing demand area under the pandemic and to make adjustment on the locations and capacity of the stations and the rebalancing schemes accordingly.

## INTRODUCTION

The outbreak of COVID-19 has seriously threatened the lives of people around the world. According to Johns Hopkins University in the United States, the cumulative number of deaths due to COVID-19 in the United States has exceeded 650,000 and the cumulative number of confirmed cases has exceeded 40.4 million as of September 8,2021. During this period, US states have implemented policies to reduce community spread of the virus, including stay-at-home orders ^[1,2]^. This order has largely impacted residents’ daily travel behavior and further affected urban transportation system. Given that the pandemic may last for long time, this impact is likely to continue.

Under the influence of the risk of exposure to COVID-19, people tend to reduce their use of public transportation such as subways and buses with closed spaces and crowded environments. However, the use of bike sharing has not been severely impacted because users could ride bikes in the open space. Studies have shown that when the public transportation system is considered dangerous during COVID-19 ^[2]^, residents usually switch from a high-risk mode to cycling to reduce the risk of infection ^[3]^. As a result, the spatiotemporal variation of demand for the use of bike sharing has changed dramatically compared with the period before the outbreak of COVID-19 ^[54]^. Therefore, understanding how the built environment factors affect the usage of public bicycles under the influence of COVID-19 is necessary because it could provide important reference to transportation planners and bike sharing operators to determine the high demand areas and to make adjustment to the location as well as the capacity of bike sharing stations.

There have been abundant works on the use of public bicycles in the period before the outbreak of COVI-19. Two types of models, namely, the global regression model ^[5,6]^ and the local regression model ^[7,8]^, are mainly used in previous studies. Global models such as linear regression model ^[7]^ and negative binomial regression model ^[9]^ assume that the coefficients of all predictors do not change across space. Although they are widely used, they cannot capture the spatial variation of the relationship between the predicting variables and the response variable, especially in the case of a large study area. Therefore, local regression models ^[7]^ such as a geographically weighted regression (GWR) model is always adopted to capture this spatial variation. Bcause the GWR model assumes that all the variables have a spatially varying relationship with the response variable, which may not be true, the semi-parametric GWR (S-GWR) model is developed to allow some variables to be global variables and other variables to be local variables.

As a result, this study sets out to explore the spatially varying relationship between the built environment and the bike sharing ridership after the outbreak of COVID-19 while controlling for the ridership before the outbreak of COVID-19. We intend to answer the following four questions.

1. Does the bike sharing ridership increase or decrease in total due to the outbreak of COVID-19?
2. If total bike sharing ridership changes, is the change of ridership of each station in proportion to the total ridership change?
3. If the change of ridership of each station is not in proportion to the total ridership change, what factors, including built environment and demographics, result in this difference?
4. How do these factors influence this difference?

The rest of the paper is structured as follows. The second part is a summary of relevant studies on the use of public bicycles. The third part describes the data used in this study. The fourth part presents the model results. The fifth part concludes this study by summarizing the main findings and the limitations.

## LITERATURE REVIEW

The literature review of related studies is composed of two parts: the influencing factors of public bicycle ridership before the outbreak of COVID-19 and the impact of COVID-19 on public bicycle usage.

### 2.1 Factors influencing public bicycle ridership before the outbreak of COVID-19

Scholars have used different data and models to explore the factors that significantly influence public bicycle ridership. The factors can be divided into two categories: external and internal.

External factors mainly refer to built environment factors, including density, diversity, and design ^[7,8,13,14]^. Sometimes, demographic factors are also regarded as the built environment factors, including age, gender, race, private car ownership, and income ^[15∼17]^. Other significant factors that are found by previous studies include weather conditions (e.g., temperature, humidity, and wind speed), weekends and holidays ^[18∼20]^. Internal factors mainly refer to personal preference and level of service. Studies have explored how the users’ willingness to use and fare influence the ridership ^[5,21∼23]^.

In terms of model selection, Rixey, Li et al., and Tran et al. used MLR models to explore the significant influencing variables ^[6,11∼12,24]^. Because the ordinary least square model cannot explain the problems of multicollinearity and spatial autocorrelation, some researchers such as Hu and Chen used partial least square to deal with the multicollinearity between variables ^[25]^. Faghih-Imani et al., Zhang et al., Wang and Chen, and Shen et al. used spatial regression models to deal with the problem of spatial autocorrelation ^[9,26∼28]^. Singhvi et al., Wang et al., and An et al. adopted the generalized linear regression models such as log linear regression to deal with the issue of skewed distribution ^[29∼31]^. Scholars such as Hu et al., Sun et al., and Hyland et al. used generalized mixed-effect model to count for the temporal autocorrelation ^[32∼34]^.

The predicting variables, response variables, and models of the most relevant studies are summarized in **Table 1**.

**TABLE 1.** Summary of studies on the relationship between built environment and bike sharing ridership.

| Author | Predicting variable |  |  |  |  |  |  |  |  |  |  |  |  |  |  | Response variable | Model |
| --- | --- | --- | --- | --- | --- | --- | --- | --- | --- | --- | --- | --- | --- | --- | --- | --- | --- |
|  | Density |  | Diversity |  | Destination accessibility |  | Distance to public transit |  |  |  | Design |  |  |  |  |  |  |
|  | Population | Employment | Mix land use | Percent of different land use types | Commute distance | Distance to the city center | Number of bus stops | Number of subway stations | Distance to the nearest bus station | Distance to the nearest subway station | Bike station characteristics | Bike lane characteristics | Road network density | Primary and secondary road density | Intersection density |  |  |
| Yang et al.<sup>[7]</sup> | ✓ | ✓ |  |  | ✓ | ✓ |  |  |  |  | ✓ |  | ✓ | ✓ |  | Hourly ridership | MLR; GWR; S-GWR |
| Ji et al.<sup>[8]</sup> | ✓ | ✓ |  | ✓ |  |  | ✓ | ✓ |  |  | ✓ |  | ✓ |  |  | Number of metro transfer trips in three weeks | GWPR |
| Faghih-Imani et al.<sup>[9]</sup> (2016a) | ✓ | ✓ |  | ✓ |  |  |  | ✓ |  |  | ✓ | ✓ |  |  |  | Hourly ridership | SLM; SEM |
| El-Assi et al.<sup>[19]</sup> | ✓ | ✓ |  | ✓ |  |  |  |  |  |  | ✓ | ✓ | ✓ |  |  | Hourly ridership | Linear mixed model |
| Lin et al.<sup>[20]</sup> |  |  |  |  |  |  |  |  |  |  |  |  |  |  |  | Annual ridership | MLR |
| Noland et al.<sup>[21]</sup> | ✓ | ✓ | ✓ | ✓ |  |  |  |  |  |  | ✓ | ✓ |  |  |  | Monthly ridership | Negative binomial regression |
| Wang and Chen<sup>[27]</sup> | ✓ | ✓ |  | ✓ |  |  | ✓ | ✓ |  |  | ✓ | ✓ |  |  | ✓ | Monthly ridership | SEM |
| Hyland M et al.<sup>[34]</sup> | ✓ | ✓ |  | ✓ |  | ✓ | ✓ | ✓ |  | ✓ | ✓ | ✓ |  |  |  | Monthly ridership | Mixed multi-layer linear model |

### 2.2 Impact of COVID-19 on public bicycle ridership

The studies related to the impact of COVID-19 on the ridership of public bicycle is summarized in this section. Buehler and Pucher studied the impact of COVID-19 on public bicycle use by means of national surveys ^[10]^. They revealed the overall trend of cycling in different cities of Europe, America, and Australia from 2019 to 2020 and the changes over time. Bucsky et al. found that the demand for public bicycles had a small decline during COVID-19 ^[4]^. A few studies have investigated factors affecting public bicycle usage under the epidemic. Hu and Chen utilized Bayesian structure time series models and partial least square regression to explore the impact of built environment and demographic variables on bike sharing ridership ^[25]^. Hu et al. used the generalized additive mixed model to explore the nonlinear relationship between influencing factors and the bike sharing ridership ^[32]^. The results showed that the number of confirmed cases of COVID-19, income level, age, and race are all significant factors.

In summary, the methods used in these studies are useful for investigating the use of public bicycles during COVID-19. Most of them used generalized linear regression models to analyze the relationship between predictors and response variables. However, the spatial nonstationarity of the relationship was not considered, which may lead to bias in the modeling results. Therefore, this paper will use GWR and S-GWR model to deal with this issue.

## DATA DESCRIPTION

This study uses two data sets, namely, the trip data of the Chicago Divvy public bicycle system and the Smart Location Database (SLD) developed by the U.S. Environmental Protection Agency. The data of the Divvy public bicycle system include start/end time of each trip, start/end station of each trip, type of membership, and user information. SLD provides the built environment and demographic information aggregated at the level of census block group (CBG).

### 3.1 Changes in the spatial distribution of usage during COVID-19

Considering that the spread of COVID-19 began on 2020/2/26 in the United States and the period of 2020/2/27 to 2020/3/4 could be regarded as the epidemic spread week, the bike sharing trip data of the eight weeks before and after the spread week are used in this study to represent the pre-COVID-19 period and post-COVID-19 period.

The weekly usage of the Divvy system in the eight weeks before and after COVID-19 is plotted in Figure 1. The usage is defined as the sum of the pickup and return trips.

**Figure 1.**
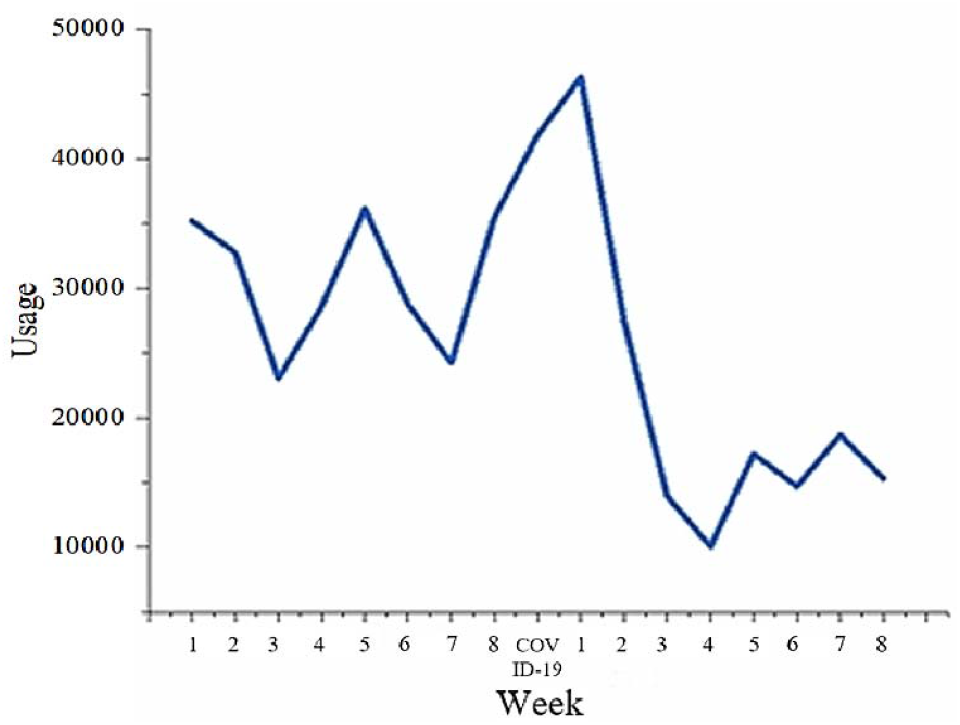
Weekly usage of bike sharing during the period of eight weeks before and after the outbreak of COVID-19.

As shown in **Figure 1**, the usage has been fluctuating around 3000 before the COVID-19. After the COVID-19, a short rise was observed, followed by a quick decline from nearly 5000 to 1000. After that, the usage became stable, being around 1800. Therefore, the research period after the epidemic in this study is from the next three weeks to the next six weeks (3-6 weeks to the right of the COVID-19 week in **Figure 1**), and the research period before the epidemic is selected symmetrically from the first six weeks to the first three weeks as the research interval (3-6 weeks to the left of the COVID-19 week in **Figure 1**). According to statistics, the average weekly usage before the epidemic was 29131.5 times, and the average weekly usage after the epidemic was 13970.25 times. Overall, the usage of public bicycles after the COVID-19 was 52.04% lower than before.

If the ratio of public bicycle usage during the COVID-19 to that before COVID-19 is roughly the same for each station, the change of usage is only caused by the epidemic and has nothing to do with other factors. If this ratio differs greatly from station to station, it shows that the change is not only affected by the COVID-19, but also affected by the characteristics of the station and surrounding environment, such as built environment and demographics. The histogram of this ratio is shown in **Figure 2**.

**Figure 2.**
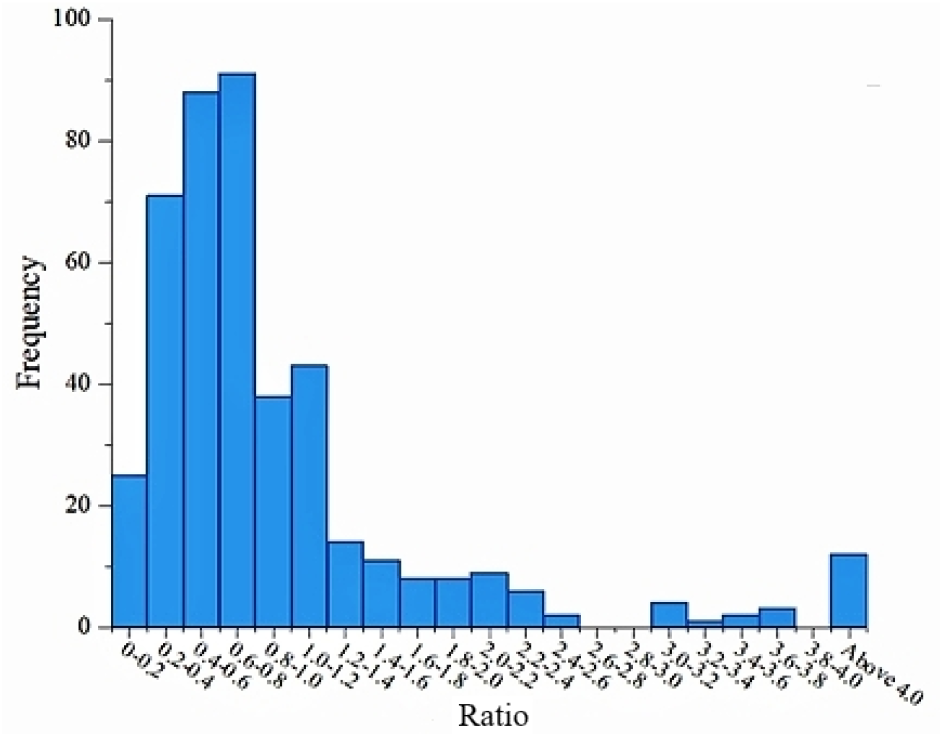
Histogram of the ratio of the ridership during COVID-19 to the ridership before COVID-19.

**Figure 2** shows that the ridership of most stations decreased during the COVID-19 while that of some stations increased and the ratio varies greatly. **Figure 3** presents the spatial distribution of the ratio. The two figures indicate that the change of usage is affected not only by the outbreak of COVID-19 but also by other factors such as the built environment.

**Figure 3.**
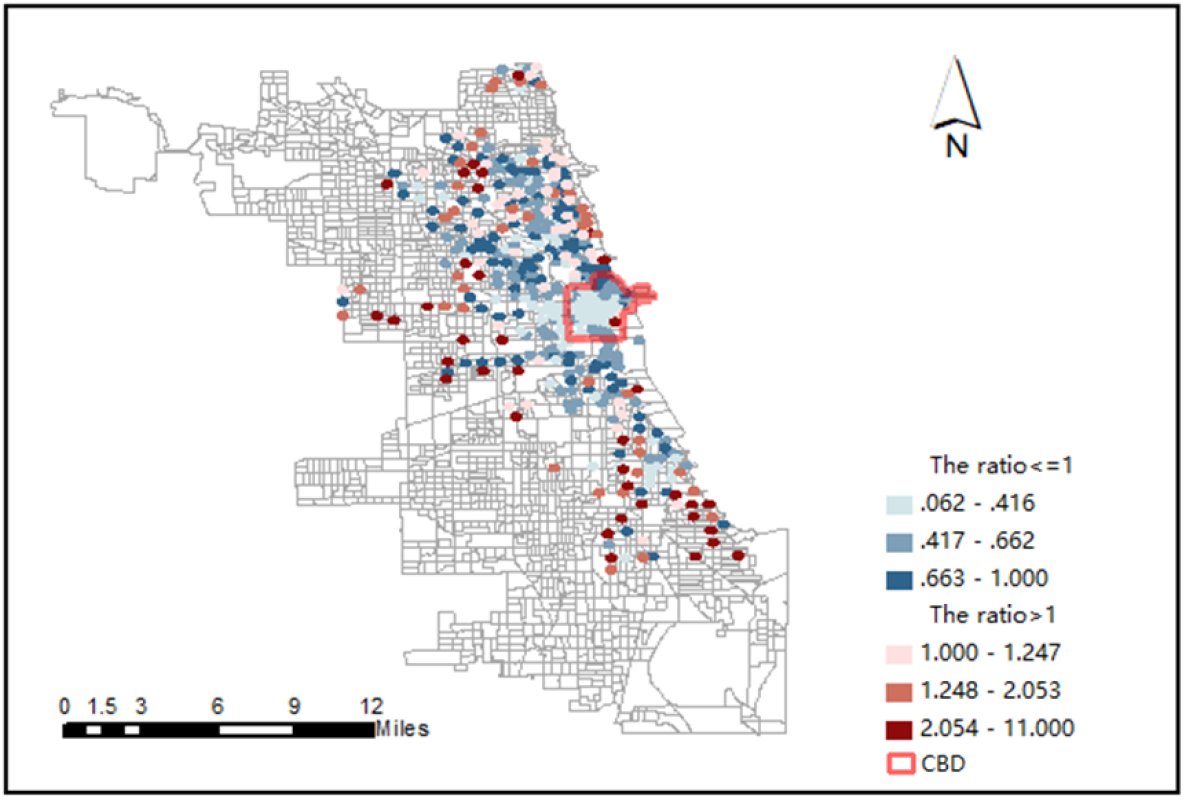
Spatial distribution of the ratio of ridership during and before COVID-19.

Figure 4 and Figure 5 present the spatial distribution of ridership. It shows that the stations with high ridership were centered around the CBD before COVID-19 and centered around the stations on the north after COVID-19. From these two figures, we can infer that bike sharing operators should pay more attention on the possible shortage of bicycles or docks of the stations on the north after the COVID-19.

**Figure 4.**
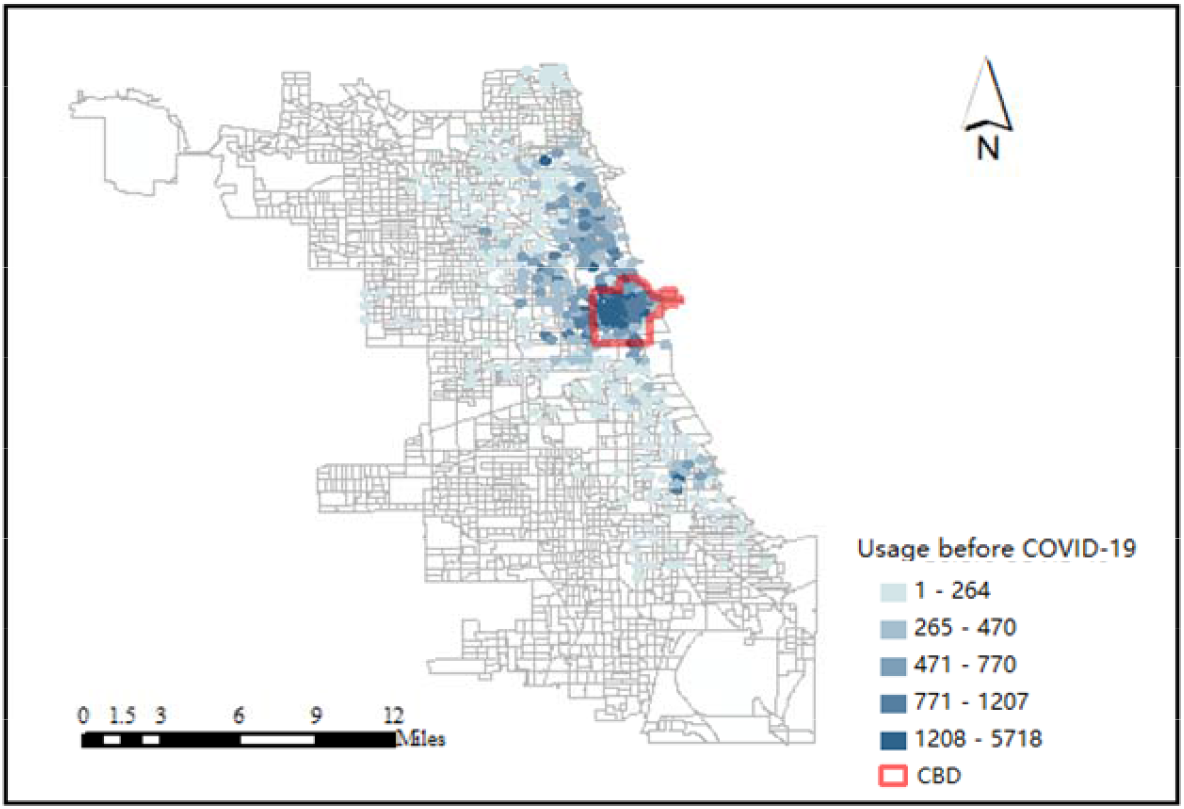
Spatial distribution of Divvy usage before COVID-19.

**Figure 5.**
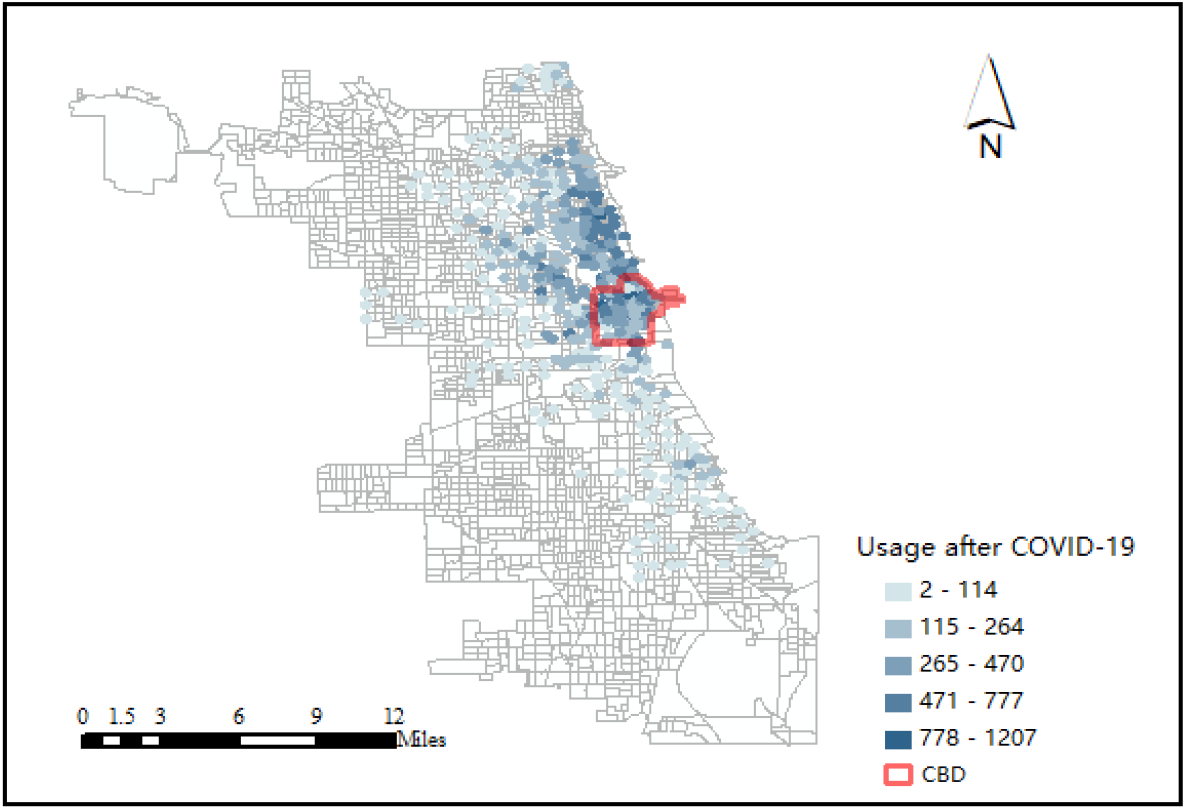
Spatial distribution of Divvy usage after COVID-19.

### 3.2 Response and predicting variables

This study used Divvy system usage in the four weeks before and after COVID-19 as the response variable.

In order to study the factors that affect the change of usage after COVID-19, it is necessary to control for the usage before COVID-19. As a result, it is included in the model as a control variable. When selecting built environment factors, this article refers to the 5D theory proposed by previous studies and related studies to select variables that may have an impact on the ridership under the COVID-19 situation ^[17]^. In the end, a total of twenty predictors including built environment, demographics, COVID-19 related cases and deaths, and ridership before COVID-19 are selected.

Seventeen variables in the built environment and demographics are derived from SLD. A circular buffer with the radius of 300 meters is drawn around each station. The radius of 300 meters is determined based on the common walking distance between the origin/destination and the public bicycle station ^[8]^. Values of the predicting variabels are extracted based on the buffer.

Number of COVID-19 cases and deaths are obtained from the City of Chicago. Considering that the COVID-19 cases and deaths may have a wider influence area ^[35]^, a circular buffer zone with a radius of 500 meters are drawn around each station to extract the number of COVID-19 cases and deaths.

### 3.3 Data processing

The histogram of the response variable shows that its distribution deviates from the normal distribution. In response to this problem, logarithmic transformation of the variable is performed. The transformed results are shown in Figure 6.

**Figure 6.**
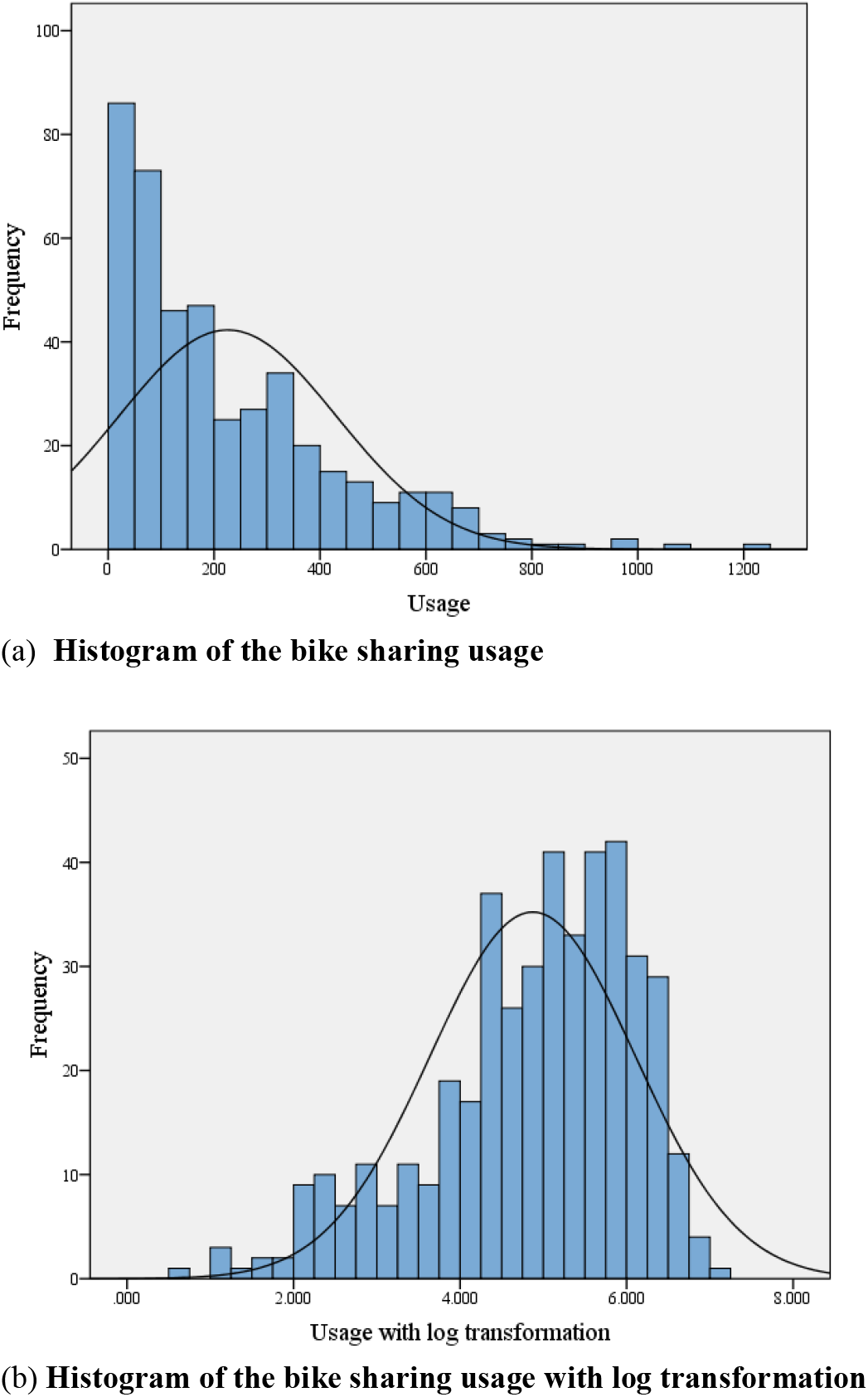
Histogram of the bike sharing usage with and without log transformation.

In order to eliminate the large difference in the magnitude of the predicting variables and facilitate result interpretation, the predicting variables are also logarithmically transformed. The modeling result will represent the elasticity of the response variable to the predicting variable, which is expressed as the percentage of change in the response variable caused by 1% change of the predicting variable. The descriptive statistics of all variables in this study are shown in **Table 2**.

**TABLE 2.**
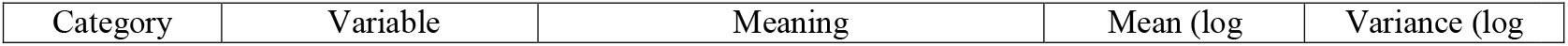
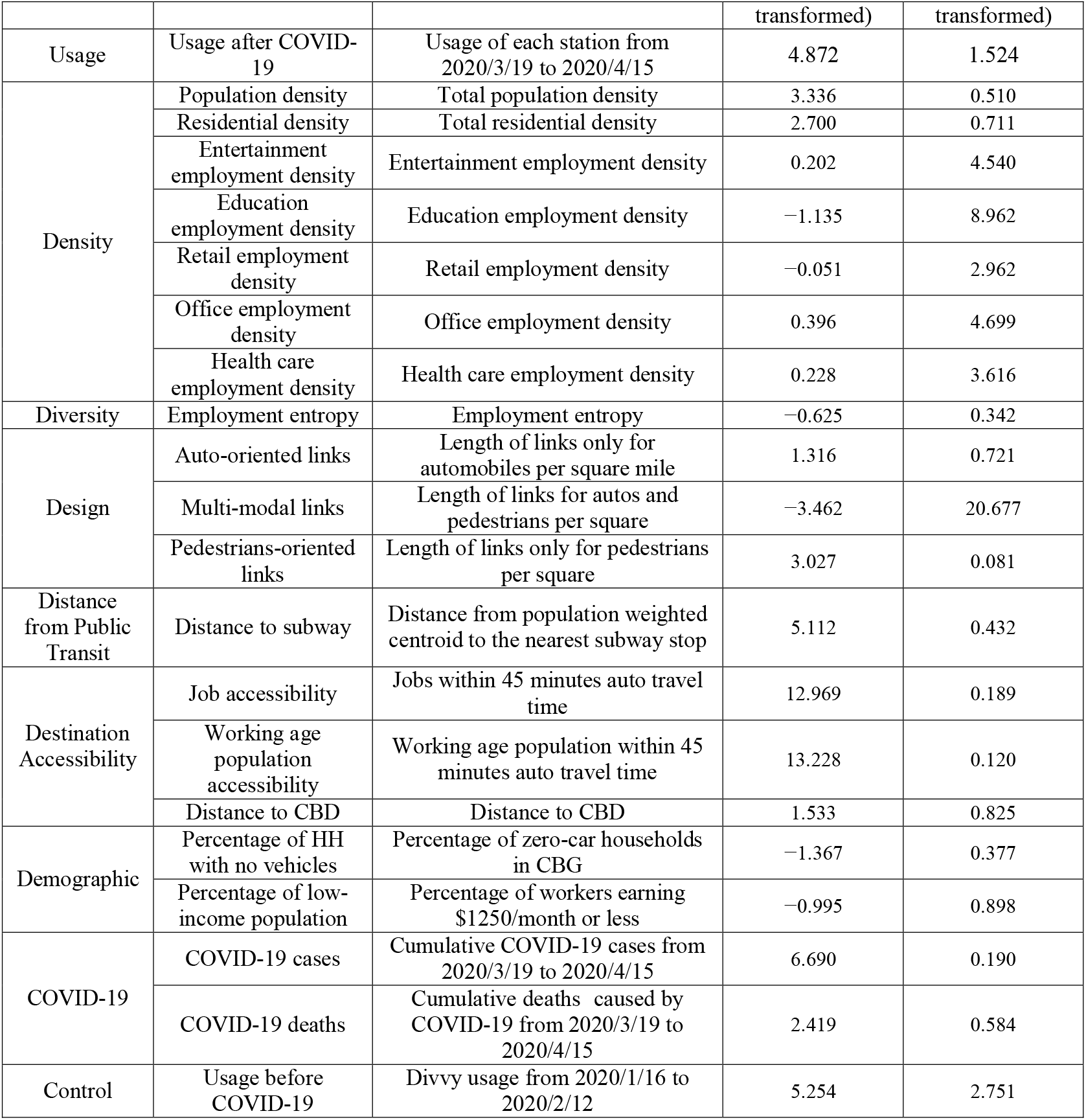
Descriptive statistics of variables.

**TABLE 3.** MLR model results.

| Variable | Unstandardized Coefficient | Significance | Collinearity statistics |
| --- | --- | --- | --- |

|  | B | Standard Error |  | VIF |
| --- | --- | --- | --- | --- |
| Residential density | 0.288 | 0.041 | .000 | 1.953 |
| Education employment density | −0.028 | 0.011 | .013 | 1.865 |
| Office employment density | −0.043 | 0.021 | .047 | 3.629 |
| Distance to subway | −0.165 | 0.047 | .000 | 1.580 |
| COVID-19 cases | 0.224 | 0.069 | .001 | 1.522 |
| Ridership before COVID-19 | 0.626 | 0.022 | .000 | 2.133 |

## 4. METHODS

### 4.1 MLR

This study uses SPSS to establish the MLR model to analyze factors that change the usage of bike sharing under COVID-19. The model assumes that the relationship between predictors and the response variable is linear and homogeneous across space. Its function is as shown in **Equation 1**.

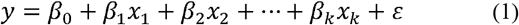

Among them, y is the response variable; x_1_, x_2_,…,x_k_ are the predictors; β_0_,β_1_,…,β_k_ are the coefficients of the predictors; ε is the random error, which has an expected value of zero, follows the normal distribution, and is independent of each other.

### 4.2 GWR

GWR model improves the traditional MLR model by allowing the relationship between predictors and the response variable to vary across space. Its function is as follows ^[7]^ (**Equation 2)**:

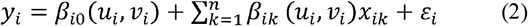

Among them, y_i_ represents the bike sharing usage of station i; β_ik_ is the coefficient of the predictor k of station i; x_ik_ is the predictor k of station i; ε_i_ is the random error term of station i; (u_i_, v_i_) represents the latitude and longitude of station i.

The predictors in the GWR model have local coefficients at each public bicycle station. When estimating the coefficients of each station, the weight w_i_ is assigned based on the distance from other stations to the target station, and the coefficient of this predictor is estimated by minimizing the weighted sum of squares. The expression is as follows (**Equation 3)**.

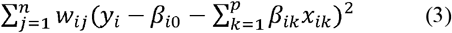

The spatial weight reflects the importance of the position. Many ways can be used to calculate the spatial weight. The simplest one is the distance threshold function. The specific function is **Equation 4**.

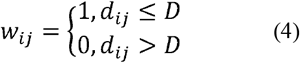

Among them, D represents the distance threshold; d_ij_ represents the distance between station i and the target station j. To solve the problem of weight discontinuity, Gaussian function is also often used to express the relationship between weight and distance (**Equation 5)**:

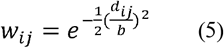

Among them, w_ij_ represents the weight between stations i and the target station j ; d_ij_ represents the distance between stations i and the target station j; b is the bandwidth.

### 4.3 S-GWR

GWR models assume that the coefficients of all predictors vary across space. However, the relationship between some predictors and the response variable may not vary across space. The S-GWR model, as an extension of the GWR model, allow some predictors to be global and others to be local. The expression of the S-GWR model is as follows ^[7]^ (**Equation 6**), and the symbols in the equation is the same as the GWR model:

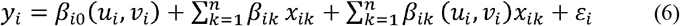

## 5. MODEL RESULTS

This section establishes MLR, GWR, and S-GWR models to explore the relationship between predictors and the response variable.

### 5.1 Results of MLR model

Backward variable selection method is adopted to select variables. Six predicting variables are significantly related to the use of Divvy public bicycles after COVID-19. These variables are residential density, education employment density, office employment density, distance to the nearest subway station, COVD-19 cumulative cases, and ridership before COVID-19.

The interpretation of the relationship between the significant predictors and the response variable is as follows:

The residential density is positively correlated with ridership after COVID-19. This may be due to the policies of work from home, which made residents to stay at home. Thus, more trips are home based trips.

The education employment density is negatively correlated. It could be due to that most schools require students and faculty members to stay at home and to take or teach classes online.

The office employment density is negatively correlated. This result may also be due to the stay at home order and work from home policy in Chicago in response to the COVID-19.

The ridership is negatively correlated with the distance to the nearest subway station. The possible reason is that residents who are close to the nearest subway station after the COVID-19 outbreak have swithed from using subway to using public bicycles.

The cumulative number of cases is positively correlated with the ridership. This result may be due to those areas with more bike sharing trips are areas with high travel demand. More people gather around the area leads to higher risk of infection. On the other hand, the increase in the number of COVID-19 infections may also make people to use public bicycles to replace other public transportation modes, such as subway and bus.

The usage of Divvy public bicycles before COVID-19 is positively correlated with the usage of Divvy public bicycles after COVID-19. This shows that stations with a high usage rate before COVID-19 continue to have high usage after COVID-19. The coefficient is 0.626, indicating that if the pre-COVID-19 usage increases by 1%, the post-COVID-19 usage will increase by 0.626% on average.

### 5.2 GWR model results

The GWR model is constructed and the coefficients of variables are shown in Table 4.

**TABLE 4.** Coefficients of each variable in the GWR model.

| Variable | Min | Lwr Quartile | Median | Upr Quartile | Max |
| --- | --- | --- | --- | --- | --- |
| Intercept | 4.650583 | 4.796953 | 5.013346 | 5.089241 | 5.200212 |
| Residential density | −0.086092 | 0.193541 | 0.277111 | 0.305725 | 0.373394 |
| Education employment density | −0.174001 | −0.086815 | −0.050423 | −0.017703 | 0.182649 |
| Office employment density | −0.245413 | −0.150852 | −0.040037 | 0.046748 | 0.177302 |
| Distance to subway | −0.286487 | −0.115407 | −0.097054 | −0.072675 | 0.084666 |
| COVID-19 cumulative cases | −0.136410 | 0.006409 | 0.035964 | 0.059660 | 0.180760 |
| Ridership before COVID-19 | 0.818073 | 0.935148 | 0.982256 | 1.030879 | 1.144220 |

The overall R-squared of the GWR model is 0.865. The R-squared of each station is visualized in **Figure 7**.

**Figure 7.**
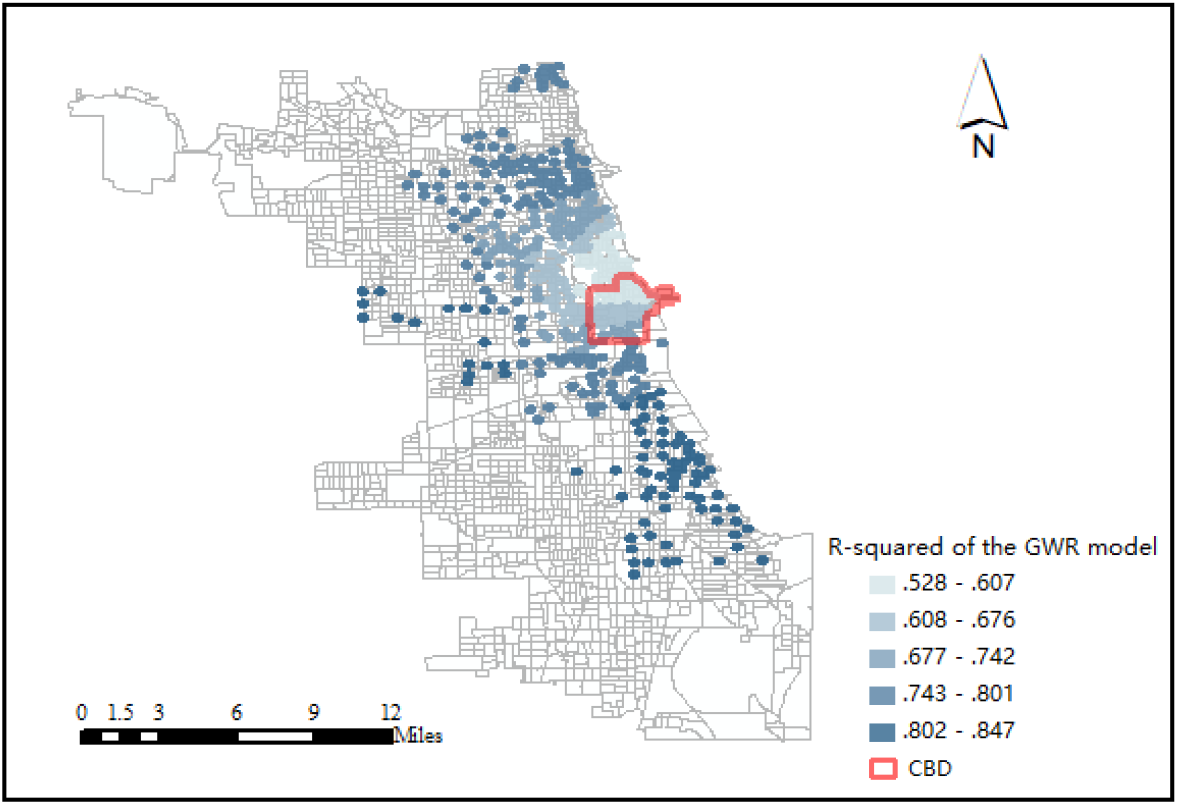
Spatial distribution of the R-squared of the GWR model.

The R-squred of each station is above 0.5 with a considerable proportion being above 0.8, which indicates that the GWR model fits the data well. The goodness of fit in the city center is generally low, which may be due to the fact that the ridership changes more in these stations than other stations probabaly due to the work from home policy, and the selected variables cannot accurately explain the dramatic change.

### 5.3 S-GWR model results

The S-GWR model is also constructed and the results of this model are shown in Table 5. To determine each predicting variable is global or local, all variables are first assumed to be global variables. The DIFF value of each variable is calculated. If DIFF>0, the predicting variable is regarded as a global variable; otherwise, it is a local variable. TABLE 5 shows that the residential density and the office employment density are local variables, and the others are global variables.

**TABLE 5.**
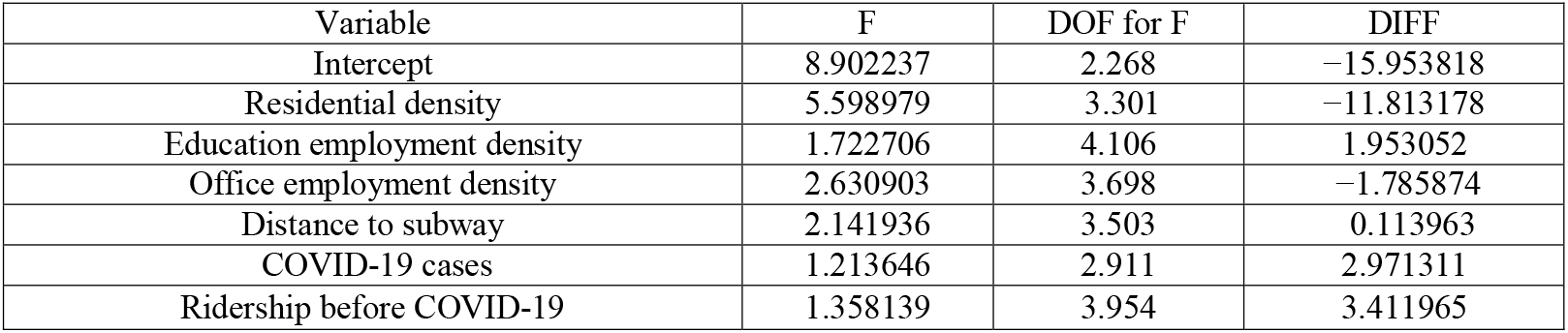
Variable type test.

| Variable | F | DOF for F | DIFF |
| --- | --- | --- | --- |
| Intercept | 8.902237 | 2.268 | -15.953818 |
| Residential density | 5.598979 | 3.301 | -11.813178 |
| Education employment density | 1.722706 | 4.106 | 1.953052 |
| Office employment density | 2.630903 | 3.698 | -1.785874 |
| Distance to subway | 2.141936 | 3.503 | 0.113963 |
| COVID-19 cases | 1.213646 | 2.911 | 2.971311 |
| Ridership before COVID-19 | 1.358139 | 3.954 | 3.411965 |

The coefficients of the two local variables will be presented below. The interpretations of the results will also be given.

The spatial distribution of the coefficients of residential density is shown in **Figure 8**. When residential density is a significant factor, it usually has a positive correlation with the usage of public bicycles after the epidemic, which is consistent with the conclusions of the previous study ^[32]^. The coefficients in the city center and the east are relatively low, which may be related to the fact that in the city center and the eastern coastal areas there are many high-income population who usually use private cars instead of public bicycles to travel. The coefficients in the southern part of the city are also relatively small. This may be due to that people living in the southern part usually have lower income. These residents may have to go out for work even after the COVID-19 outbreak. Thus, the number of home-based trips per person is low.

**Figure 8.**
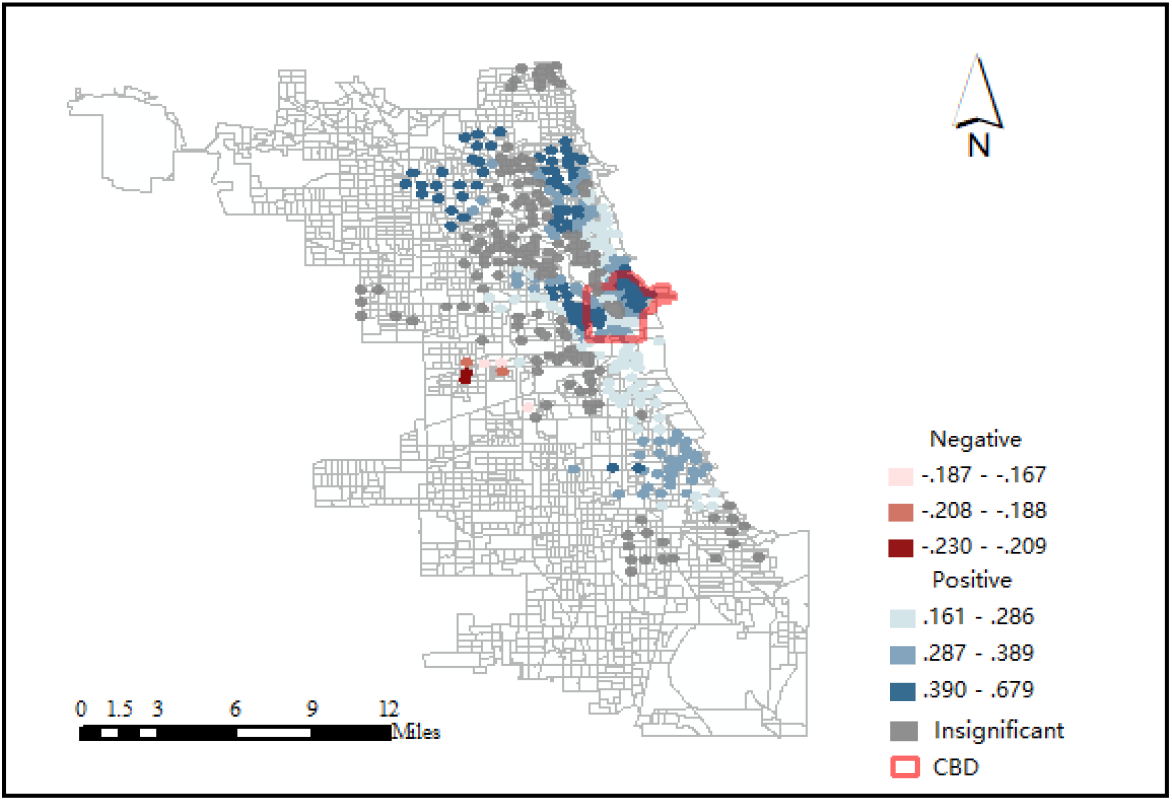
Distribution of residential density regression coefficients.

The spatial distribution of the coefficients of office employment density is shown in **Figure 9**. When the office employment density is a significant factor, it is usualy negatively correlated with the ridership. It is probably because the stay-at-home order during COVID-19 reduced the number of people working in the office and thus reduced the usage of public bicycles in the areas with high office employment density. Therefore, this negative impact is very obvious in the city center.

**Figure 9.**
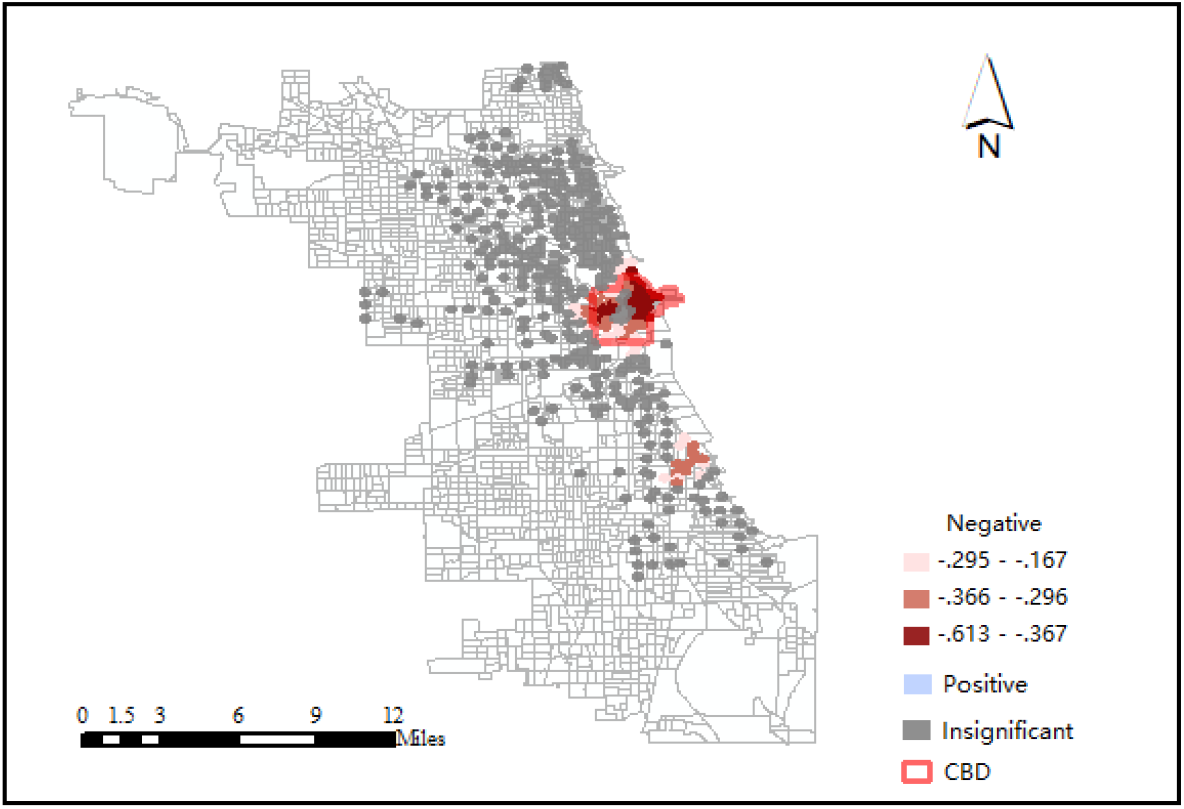
Distribution of office employment density regression coefficients.

The overall R-squared of the S-GWR model is 0.886, which is higher than that of the GWR model. The spatial distribution of the R-squared of the S-GWR model is shown in Figure 10. The goodness-of-fit of the stations in the city center is still not high, which shows that the dramatic change of ridership in these areas caused by the COVID-19 is difficult to be captured.

**Figure 10.**
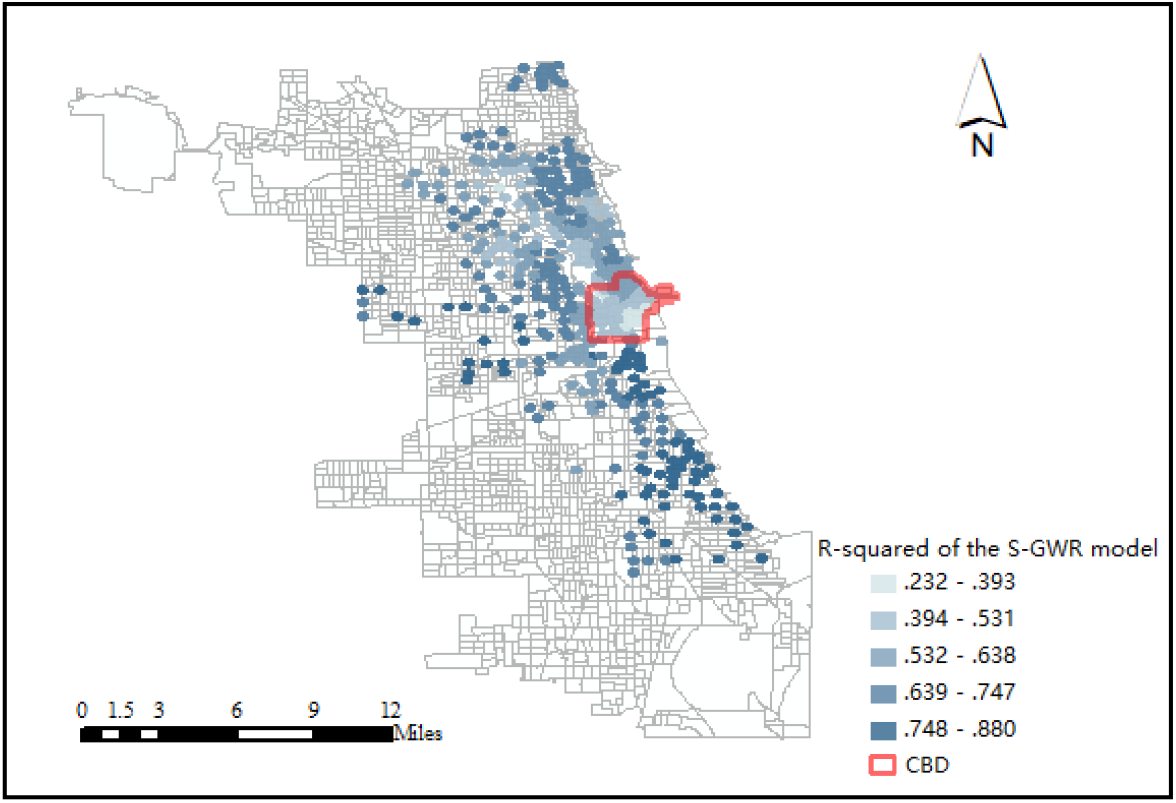
Spatial distribution of the R-squared of the S-GWR model.

### 5.4 Model comparison

The results of the MLR, GWR, and S-GWR models are compared and shown in **TABLE 6**.

**TABLE 6.** Comparison of model results.

| Measures | MLR | GWR | S-GWR |
| --- | --- | --- | --- |
| Residual sum of squares | 112.160 | 89.645 | 75.429 |
| -2 log-likelihood | 645.349 | 547.657 | 472.374 |
| Classic AIC | 661.349 | 612.292 | 591.082 |
| AICc | 661.687 | 617.640 | 610.155 |
| R-squared | 0.831 | 0.865 | 0.886 |
| Adjusted R-squared | 0.828 | 0.851 | 0.862 |

Since the models with smaller sum of squares of residuals, -2 log-likelihood, AIC, AICc, and higher-squared is regarded as better model, from these perspectives, S-GWR model is the best among the three models.

## 6. CONCLUSION

This study investigates the built environment factors that influence the bike sharing ridership of the Chicago Divvy system during the period of COVID-19 while controlling for the ridership before COVID-19. To capture the spatially varying relationship between built environment and ridership, GWR and S-GWR models are established and their results are compared to those of MLR model. We found that S-GWR has the highest goodness-of-fit from many perspectives.

We also found that the total bike sharing ridership declined by half after the outbreak of COVID-19. The decline of ridership of each station is very different and the spatial distribution of usage of bike sharing during the period of COVID-19 is very different from that before COVID-19. It indicates that transportation planners and bike sharing operators should pay attention to this change and could adjust the capacity and location of the stations as well as the rebalancing scheme according to the current ridership pattern.

In terms of the relationship between built environment and change of ridership, some variables are local variables such as residential density and office employment density and some variables are global variables such as education employment density and distance to the nearest subway station. The complex relationship should be fully considered when estimating the change of ridership of bike sharing stations.

We have to admit that there are also some limitations in this study. First of all, although the analysis framework could be applied to other cities, the results obtained from this study may not be the same as other cities. Each city should develop policies based it own condition. Secondly, because we used the cross-sectional data, the revealed relationship between the independent variables and the response variable should be regarded as correlation instead of causal relationship. Although some causal relationship could be inferred from the results, this inference should be made with causion. In the future, panel data could be used to deal with this issue.

## Data Availability

These datasets were derived from the following public domain resources.
https://www.divvybikes.com/system-data https://www.epa.gov/smartgrowth/smart-location-mapping
https://data.cityofchicago.org/Health-Human-Services/COVID-19-Cases-Tests-and-Deaths-by-ZIP-Code/yhhz-zm2v
https://data.cityofchicago.org/browse?tags=gis

## ACKNOWLEDGMENTS

This study was funded by the National Natural Science Foundation of China (Grant numbers 51774241 and 71704145), China Postdoctoral Science Foundation, and Sichuan Youth Science and Technology Innovation Research Team Project (Grant numbers 2019JDTD0002 and 2020JDTD0027).

## AUTHOR CONTRIBUTIONS

The authors confirm contribution to the paper as follows. Hongtai Yang: conceptualization, methodology, formal analysis, and writing, particularly review and editing. Zishuo Guo: data processing, formal analysis, and writing, particularly original draft preparation. All authors reviewed the results and approved the final version of the manuscript.

